# Plasma concentrations of curcumin in individuals using curcumin with adjuvants or lipid formulated curcumin supplements: a real world cohort

**DOI:** 10.1101/2023.03.23.23287619

**Authors:** Maurice A.G.M. Kroon, Jacqueline K. Berbee, Soumia Majait, Noortje E.L. Swart, Olaf van Tellingen, Hanneke W.M van Laarhoven, E. Marleen Kemper

## Abstract

The spice curcumin and its metabolites have been linked to many beneficial health effects. These effects have, thus far, not been duplicated in independent research most likely due to low plasma concentrations of curcumin. Despite the many reports, the public’s interest in curcumin continues to grow and many people use curcumin in daily life. Moreover, companies seize the popularity of curcumin and claim that their formulations increase systemic expose of curcumin. In this independent study we determined the plasma concentration of curcumin after oral intake in daily life to determine if the systemic exposure is sufficient to achieve beneficial health effects.

We used a validated HPLC-MS/MS assay to determine the plasma concentration of curcumin and its metabolites in 47 individuals (patient or healthy) using their own curcumin formulations. Through questionnaires, we assessed which other supplements and (self-)medication(s) were used. The concentrations of curcumin and its metabolites were analyzed in plasma samples collected just before and 1.5 h after curcumin intake. Each sample was pretreated with and without β-glucuronidase to determine the levels of conjugated and unconjugated curcumin.

After oral intake of the curcumin supplement, plasma concentrations of curcumin, demethoxycurcumin, bisdemethoxycurcumin and tetrahydrocurcumin ranged between 2 and 4 nM. Use of adjuvants like piperine did not result in higher curcumin plasma concentrations. Adding β-glucuronidase to the plasma sample increased curcumin plasma levels from below LLQ to 25.3 ng/mL, however still below any plasma concentration to which a beneficial health effect can be expected.

The observed plasma concentration of unchanged curcumin remained several orders below the concentration of 2-100 μg/mL used in *in vitro* studies. Therefore, our study confirms the low plasma levels of curcumin and indicates the need to be critical towards the claimed beneficial systemic health effects of current curcumin supplement use in daily life among patients and healthy individuals.

## 1. Introduction

The spice curcumin originates from the root of the *Curcuma Longa* and has been used for centuries in Asian cultures as dietary pigment, spice and traditional medicine. Curcumin has been associated with many beneficial health outcomes, such as anti-inflammatory and anti-cancer effects. (1-3) However these effects have mostly been assessed in *in vitro* experiments. For example, curcumin showed activity against a human pancreatic carcinoma cell-line with IC90 values of 2.5-35 μg/L (4) and reduced the tumor growth in a mouse model for pancreatic cancer. (5) Clinical trials, however, could not confirm these results. (6, 7) One study described some beneficial clinical effect in two patients but it is questionable if this effect can be contributed to the sole use of curcumin. (8)

One of the major known issues potentially explaining the low therapeutic effectivity of curcumin in patients is the low bioavailability after oral administration. Curcumin dosages of up to 12 g show no detectable levels of curcumin in the systemic circulation. (9) The low solubility, low dissolution rate and instability in intestinal pH of curcumin will not favor absorption from the small intestine. (10) Furthermore, curcumin has been found to be subject to extensive phase II metabolism. (11) In a study with rat and human liver hepatocytes curcumin was first reduced to hexahydrocurcumin and then to hexahydrocurcuminol, whereas conjugation of curcumin is only a minor hepatic biotransformation route. (11) However, despite these findings, high levels of glucuronidated and sulfated levels of curcumin have been quantified in human plasma suggesting a high phase II metabolism via urididne-5’-diphosphoglucuronic acid (UGTs) in the gut wall. One study found that human intestine mircrosomes had about a three-fold higher activity than human liver microsomes for curcumin and demethoxycurcumin, indicating a major first pass metabolism of curcumin in the gastrointestinal tract. (12)

To overcome the low bioavailability, different curcumin formulations – like lipid formulations – have been developed. Several of these products are currently highly advertised and used by patients. (13) There has been an increase in the use of curcumin loaded liposomes, micelles, colloidal suspensions and lipid-based nanoparticles resulting in multiple clinical trials on curcumin nanoformulations registered currently on clinicaltrials.gov. Furthermore, it has been described that the addition of certain adjuvants could overcome the low bioavailability of curcumin. One clinical study showed that addition of 20 mg/kg piperine increased bioavailability of curcumin with respectively 154% in rats and 2000% in humans. (14) Piperine is suggested to inhibit the phase I and II metabolism by inhibition of CYP3A4 and inhibition of UDP-glucuronosyl transferase (UGT) and thereby inhibiting the formation of the more water soluble and excretable metabolites of curcumin thus increasing the bioavailability of curcumin. (15-17)

Despite the disappointing results in clinical studies and the knowledge of the low bioavailability, the public’s interest continues to grow in curcumin due to new formulations being developed by various manufactures. If we can assess the plasma concentration of curcumin formulations being used in daily life and differentiate between conjugated and unconjugated curcumin and its metabolites, we can better inform patients and healthy individuals about the supposed claimed beneficial effects of the current curcumin supplement use. In this independent, observational study we therefore addressed the following questions: 1. What is the plasma concentration of curcumin and the metabolites demethoxycurcumin, bisdemethoxycurcumin, tetrahydrocurcumin when being used in daily life? 2. Which adjuvants, such as piperine, do participants use to increase the bioavailability of curcumin.

## 2. Method

### 2.1 Study participants

All participants were recruited through free advertisements placed inside the Amsterdam UMC, location AMC Hospital from 1^st^ of December 2016 till 21^st^ September 2017. Forty-seven adult persons (patients or healthy volunteers) signed the informed consent. The inclusion criteria comprised of age 18 years or older, mentally competent to give informed consent and use of curcumin (food) supplement in daily life. Participants were asked to give information about their health condition and provided, where possible, an overview of their prescribed drugs. Un-prescribed medication, use of food supplements and/or other herbal drugs were also recorded. Participants were allowed to participate twice – after a minimal three day washout period – with different curcumin formulations. The study protocol was reviewed and approved by the Ethics committee of the Amsterdam UMC, location Academic Medical Centre, The Netherlands. The study was registered in the Dutch Clinical Trial Register with ID NL5931.

### 2.2 Study design

This study is an observational, open-label study investigating concentration of curcumin as used in daily life. Each participant was instructed to bring their own curcumin formulation on the day of visit. Participants were asked not to take their curcumin formulation in the morning on the day of visit. Blood was taken just before intake of the curcumin supplement (trough concentration) and 1.5 hours after intake (expected peak concentration). Any food or beverage intake between these time points was registered while participants were not allowed to take any additional curcumin in between these two time points.

### 2.3 Blood sampling and processing

Blood was collected in heparinized tubes and protected against light by wrapping in aluminum foil. All samples were centrifuged (Rotina 380R from Hettich) for 10 min at 2754 RCF and 20°C and the obtained plasma was stored, protected from light, at -20 °C until analysis.

### 2.4 Analysis of curcumin, demethoxycurcumin, bisdemethoxycurcumin, tetrahydrocurcumin and piperine in plasma

Plasma samples were analyzed by our HPLC-MS/MS method (18). In short, on the day of analysis, samples were thawed and pretreated with and without β-glucoronidase. The samples receiving treatment with β-glucoronidase were incubated with 20 U/μL β-glucuronidase at 37°C for 1 hour. Liquid-liquid extraction with Tert Butyl Methyl Ether (TBME) as organic phase was used to extract the compounds from the plasma. Curcumin, demethoxycurcumin, bisdemethoxycurcumin, tetrahydrocurcumin and piperine were quantified in a single analytical run using a validated HPLC-MS/MS method. The HPLC-MS/MS system consisted of an Ultimate 3000 autosampler and pump, both of Dionex, connected to a degasser from LC Packings. The autosampler, with a 100μL sample loop, was coupled to a Sciex API4000 mass spectrometer. After injection of 50 μL sample, separation of the analytes was performed with an Agilent column 2.1×100 mm packed with material of Zorbax Extend 3.5 μm C-18. The flowrate was 0.200 μL/min and the dual gradient mobile phase consisted of A: ultra-purified H_2_O with 0.1% formic acid and B: MeOH 100%. The applied gradient profile started at 50:50 A:B and increased linear to 95% B in 3.0 minutes. During 6 minutes a 5:95 A:B level is continued, after which it returned in 0.2 minutes to 50:50. Afterwards the system was equilibrated during 6 minutes at the starting level. Throughout the liquid-liquid extraction and HPLC-MS/MS the potential influence of light was minimized by working in an dark environment as much as possible. Each analytical run included a set of freshly prepared calibration samples containing all compounds in the validated range of 2 nM to 400 nM.

### 2.5 Statistical Analysis

This is an observational study and the main aim is to assess if curcumin is detectable in the plasma of participants after oral curcumin intake. Due to the descriptive nature of this design with different dosages and different formulations, no statistical analysis was performed. Any curcumin plasma level below the LLQ of 2 nM is not expected to have any health effect. We aimed to include 50 participants to have an adequate indication of the different products used and provide preliminary evidence of the plasma curcumin levels reached when curcumin is taken in daily life.

## 3. Results

### 3.1 Participants and overview of curcumin use

Within 10 months, 47 adult persons were included, 20 males and 27 females (see figure 1). Three persons participated twice using different curcumin formulations and the interval between their participation was 3 days, 13 days and 31 days, respectively. Among the 47 participants, 170 different food supplements and 95 different prescribed drugs were reported. The food supplements can be divided into eight different categories: vitamins and minerals, herbs and other botanicals, enzymes and antioxidants, fish oil and derivatives, cannabis oil, amino acids, sedatives and miscellaneous. Vitamins and minerals were most frequently reported, followed by herbs and other botanicals. We assessed which herbal products or supplements were used that might influence the bioavailability of curcumin. Piperine, genistein or drugs metabolized by the same enzymes as curcumin, namely CYP3A4, CYP2C9, UGT and SULT have a potential effect on influence the bioavailability of curcumin. (22) Table 1 describes prescribed drugs used by participants and are divided into different drug classes. Antihypertension drugs were the most prescribed drug class, followed by corticosteroids.

**Figure 1.**
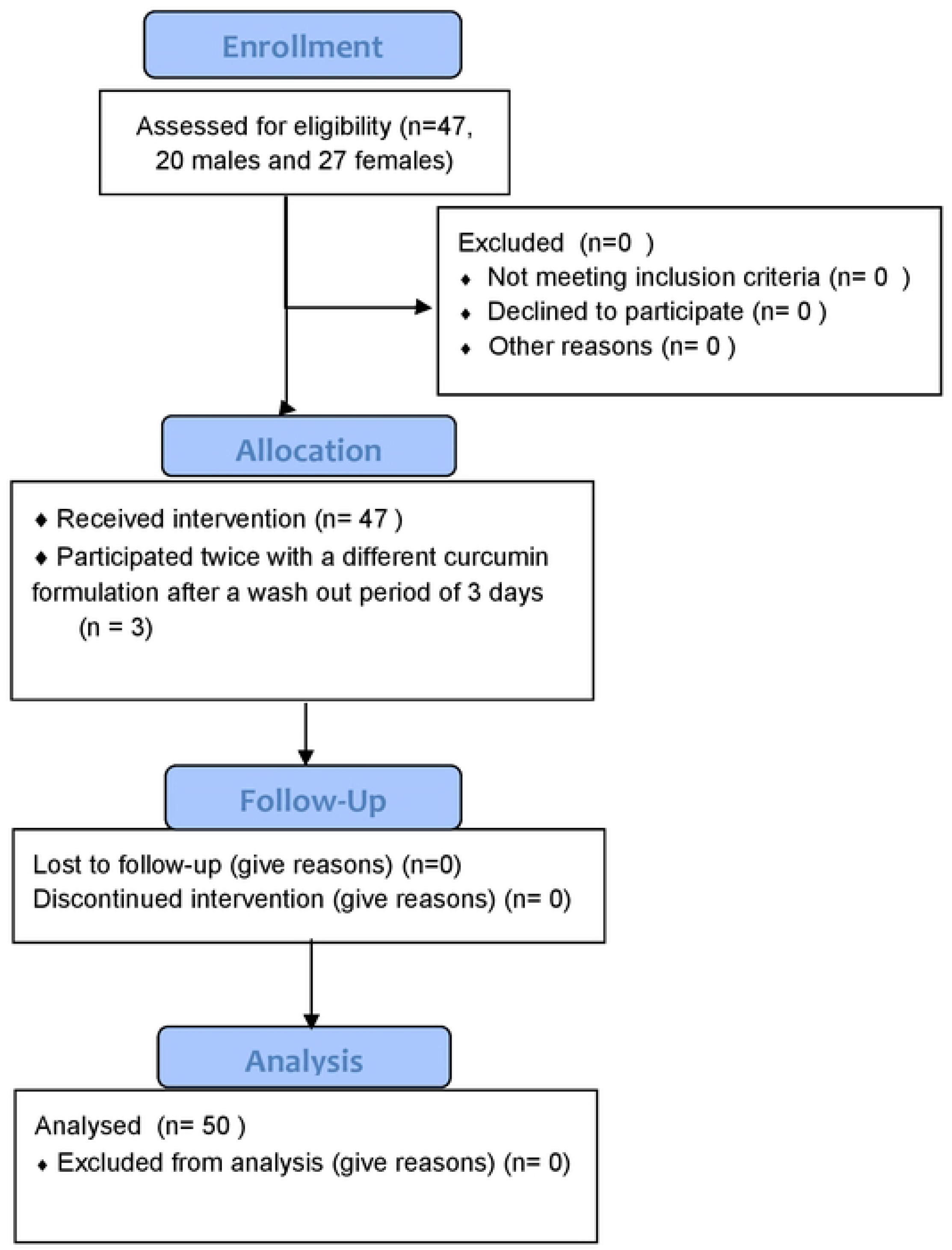
CONSERT Flowchart of enrollment, allocation, follow up and analysis. Forty-seven participants (20 males and 27 females) were included of which 3 participants participated twice resulting in 50 data points.

**Table 1.** Overview of subclasses of prescribed drugs amongst the 47 participants.

| DRUG CLASSES | FREQUENCY |
| --- | --- |
| ANTIHYPERTENSION/BETABLOCKERS | 13 |
| CORTICOSTEROIDS | 7 |
| PROTON PUMP INHIBITORS | 5 |
| VITAMINS | 5 |
| THYROID DRUGS | 4 |
| ANTICOAGULANTS | 4 |
| CHOLESTEROL LOWERING DRUGS | 3 |
| PAINKILLERS | 3 |
| ANTIDIABETIC | 3 |
| HORMONES | 3 |
| RENAL | 3 |
| ALLERGY | 3 |
| ANTI-EMETICUM | 3 |
| ANTI-DIURETIC | 2 |
| ANTIBODIES | 2 |
| OTC | 2 |
| PSYCHOPHARMACA<br>(SSRI/BENZODIAZEPINE) | 3 |
| BIRTH CONTROL | 1 |
| ANTI-EPILEPTIC | 1 |
| CYTOSTATIC | 1 |
| ENZYMES | 1 |
| ANTIVIRAL | 1 |
| LAXATIVES | 1 |
| TOTAL | 74 |

The 47 participants used in total 34 different curcumin formulations. Sixteen participants used curcumin capsules without adjuvants, whereas 22 participants used curcumin in combination with an adjuvant. Three participants used a curcumin formulation where curcumin micelles were formulated in soft gel capsule. Four participants made their own curcumin formulation (i.e. tea or in food). Five participants made their own formulations with adding adjuvants like piperine (figure 2).

**Figure 2.**
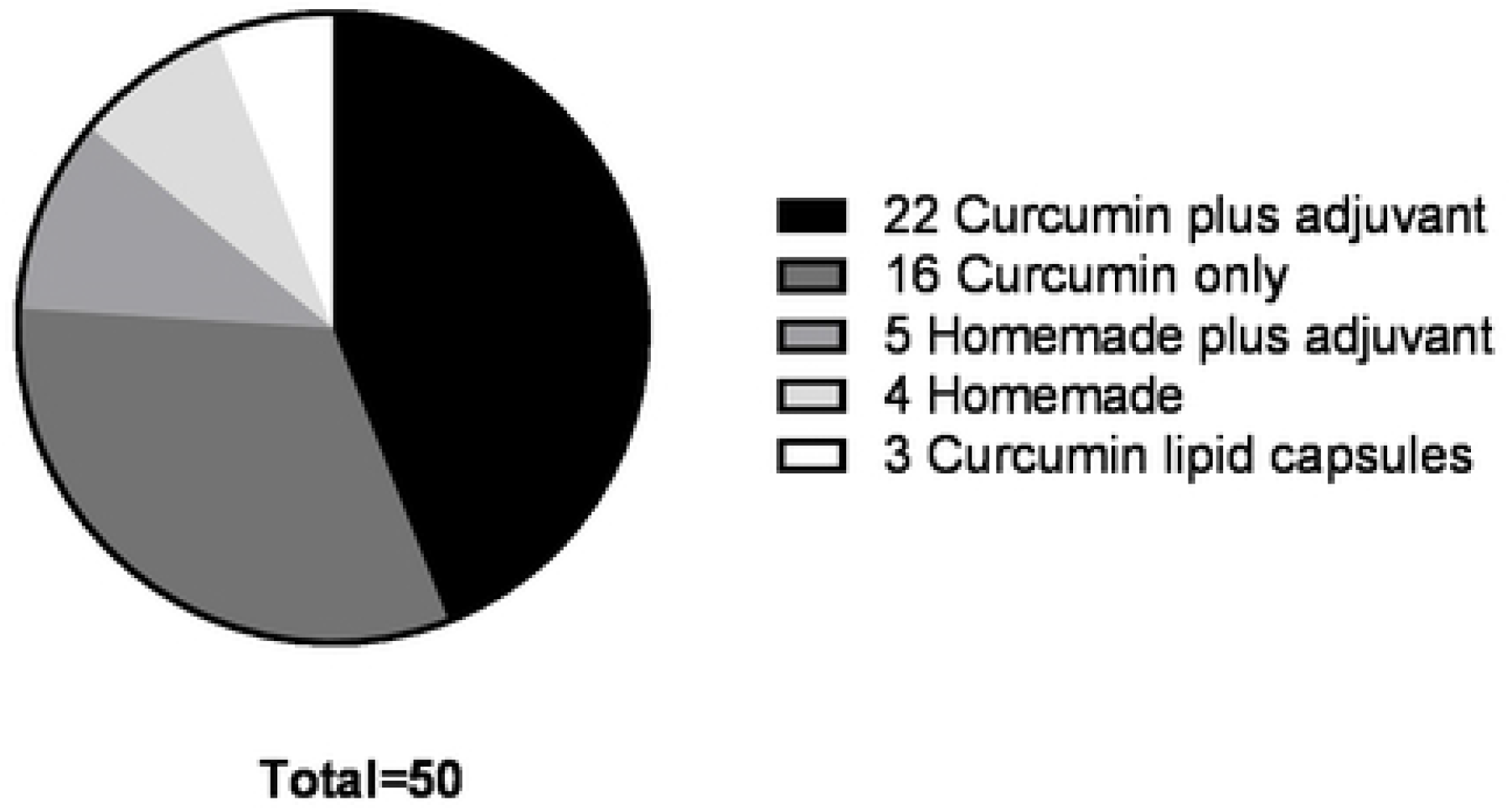
Different formulations of curcumin used among the 47participants totaling in 50 data points. Twenty-two participants used curcumin plus an adjuvant of which 22 formulations consisted of different brands curcumin capsules with added piperine and different dosages. Sixteen participants used curcumin only supplements of which 16 formulations consisted of different curcumin capsule brands in different dosages. Five participants used homemade formulations plus an adjuvant of which everybody used piperine as adjuvant. Four participants used a homemade formulation consisting of crude curcumin powder. Three participants used curcumin only in a lipid capsule formulation.

### 3.2 Plasma levels of curcumin, demethoxycurcumin, bisdemethoxycurcumin, tetrahydrocurcumin and piperine plus conjugates

All plasma samples were analyzed with and without the addition of β-glucuronidase to determine unconjugated and conjugated curcumin. The analysis of curcumin, demethoxycurcumin, bisdemethoxycurcumin, tetrahydrocurcumin and piperine of the 47 participants are shown in table 2. In only one of the 50 data points, curcumin was detected at the through level at a concentration of 10.1 ng/mL. At the expected peak levels, 1,5 h after intake, curcumin was detectable in 2 participants at 0.9 ng/mL and 1.1 ng/mL respectively. Demethoxycurcumin plasma levels were below our lowest limit of quantification (LLQ) of 0.68 ng/mL before and after intake of curcumin. Bisdemethoxycurcumin was detected in 1 person at 2.1 ng/mL at the trough level whereas this metabolite was below the LLQ at the expected peak level. Tetrahydrocurcumin was below the LLQ of 0.74 ng/mL at trough levels and only quantified in two participants after intake at 16.4 ng/mL and 20.9 ng/mL.

**Table 2.** Results of curcumin, DMC, BMC, THC and PIP in their conjugated and unconjugated forms in human plasma at through and expected peak level. All compounds are shown before and after the addition of β-glucuronidase which leads to unconjugated forms of curcumin. *n*.*d*.= not detectable (below the LLQ).

| Compound (LLQ in<br>ng/mL) | Time of<br>intake<br>(hour) | Without addition of<br>β-glucuronidase | | After addition of<br>β-glucuronidase | |
| --- | --- | --- | --- | --- | --- |
|  |  | Detectable<br>levels in #<br>persons | Mean +<br>Concentration<br>range (ng/mL) | Detectable<br>levels in #<br>persons | Mean +<br>Concentration range<br>(ng/mL) |
| **Curcumin**<br>**(0.74 ng/mL)** | 0 | 1 | 10.1 | 27 | 14.3 (0.8 – 66.3)) |
|  | 1.5 | 2 | 1.0 (0.9 – 1.1) | 42 | 25.4 (0.7 – 130.8) |
| **Demethoxycurcumin**<br>**(0.68 ng/mL)** | 0 | 0 | *n.d.* | 9 | 3.5 (1.4 – 8.6) |
|  | 1.5 | 0 | *n.d.* | 26 | 9.4 (0.9 – 73.1) |
| **Bisdemethoxycurcumin**<br>**(0.62 ng/mL)** | 0 | 1 | 2.1 | 8 | 1.6 (0.7 – 2.7) |
|  | 1.5 | 0 | *n.d.* | 25 | 6.1 (0.7 – 60.4) |
| **Tetrahydrocurcumin**<br>**(0.74 ng/mL)** | 0 | 0 | *n.d.* | 30 | 14.6 (0.8 – 81.2) |
|  | 1.5 | 2 | 18.6 (16.4 – 20.9) | 39 | 21.6 (1.0 – 161.6) |

Pretreating the samples with β-glucuronidase resulted in higher curcumin plasma concentrations the participants. After treatment with β-glucuronidase, 27 participants showed unconjugated curcumin levels at trough levels varying from 0.78 ng/mL to 66.3 ng/mL. Whereas after treatment with β-glucuronidase, unconjugated curcumin was found in 42 participants at 1.5 h after dosing (expected peak level) with concentrations varying from 0.7 ng/mL to 130.8 ng/mL. The three participants using curcumin lipid formulations showed unconjugated plasma concentrations of 80.7 ng/mL to 130.8 ng/mL and were consequently the highest found unconjugated curcumin plasma levels after the addition of β-glucuronidase. Unconjugated curcumin, demethoxycurcumin, bisdemethoxycurcumin and tetrahydrocurcumin plasma levels without the addition of β-glucuronidase, remained below the LLQ for all participants. Unconjugated demethoxycurcumin peak level concentrations was detected in 26 persons with varying concentrations of 0.9 ng/mL to 73.1 ng/mL. Unconjugated bisdemethoxycurcumin peak level concentrations were detected in 25 persons with concentrations varying from 0.7 ng/mL to 60.4 ng/mL. Unconjugated tetrahydrocurcumin peak level concentrations were detected in 39 persons with concentrations varying from 1.0 ng/mL to 161.6 ng/mL. Piperine trough and peak level concentrations were also assessed and ranged from 1.0 ng/mL to 1096 ng/mL and were detectable in all participants whether or not it was purposely used as adjuvant to curcumin.

## 4. Discussion

Our study shows that there is very little to no systemic exposure of curcumin even after use of curcumin on a continuous basis in everyday life. Adjuvants or use of different formulations did not show therapeutic concentrations of curcumin in plasma. Our study indicates the need to be critical towards the claimed beneficial effects of curcumin supplement use in daily life among patients and healthy subjects. (19)

Our first aim was to assess the plasma concentration of curcumin, demethoxycurcumin, bisdemethoxycurcumin, tetrahydrocurcumin and piperine. The in *vitro* results that have been observed throughout the literature show effective curcumin concentrations between 2-100 μg/mL. Our results show that unconjugated curcumin does not achieve concentrations above 0.74 ng/mL in the plasma of our participants which is more than a 1000-fold lower than the highest concentration assessed *in vitro* which is suspected of having beneficial health effects. Everyday use of the currently available curcumin formulations in people seem not sufficient to achieve any systemic effects.

Our validated, HPLC-MS/MS method was able to assess plasma levels in unconjugated (free curcumin) form in patients or healthy persons who use curcumin supplements in everyday life. Addition of β-glucuronidase allows us to indirectly measure conjugated (glucuronidated and sulfated forms) curcumin, demethoxycurcumin, bisdemethoxycurcumin and tetrahydrocurcumin. Our results are in contrast with results of several studies in which high systemic concentrations of curcumin are claimed. (20-22) All of these studies reported the curcumin as unconjugated plus conjugated curcumin. Our method of analysis contained a crucial step in which each plasma sample was treated with and without β-glucuronidase differentiating between unconjugated and conjugated curcumin. The β-glucuronidase enzyme hydrolyzes the hydrophilic glucuronic and sulfatic acid group from curcumin (figure 3). Analyzing the plasma sample after the addition of β-glucuronidase showed curcumin plasma levels in 42 participants. Curcumin plasma levels assessed without the use of β-glucuronidase showed plasma levels of curcumin in only two participants. This would suggest that curcumin is conjugated instantly after absorption thus showing a virtually complete first pass metabolism.

**Figure 3.**
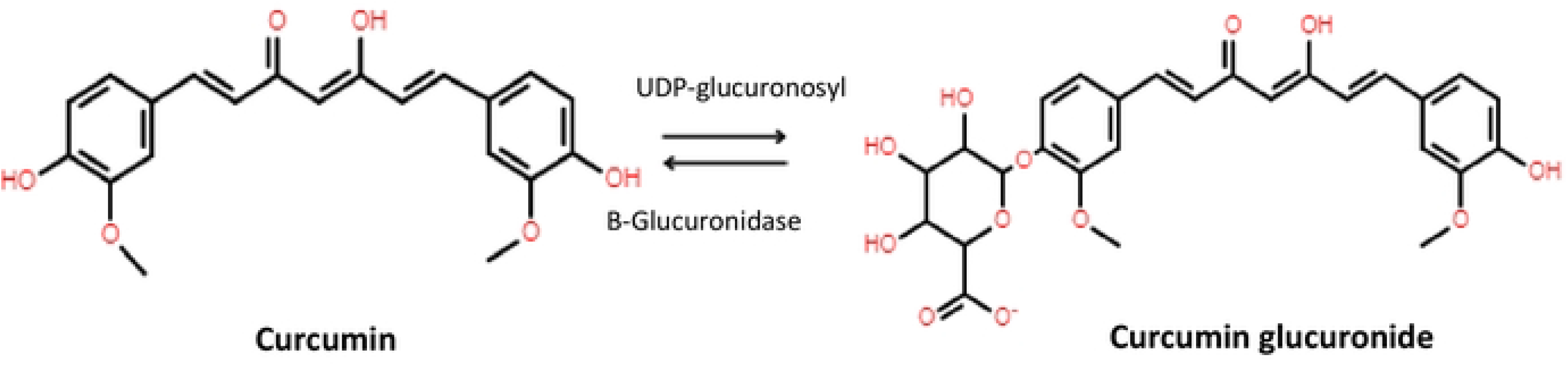
Conversion of curcumin by UDP-glucuronidase to curcumin glucuronide and back to curcumin by β-glucuronidase.

Our second aim was to determine which adjuvants, such as piperine, or supplements are used by individuals to influence the bioavailability of curcumin. It is suggested that piperine affects curcumin plasma levels at IC_50_ value on CYP3A at 5.5 ± 0.7 μM with CYP2C9, CYP2D6, CYP2C19 CYP1A2, CYP2E1 and CYP2B6 IC_50_ values to be between 29.8 to ≥50 μM. (23) Twenty-five of the 47 participants used piperine as adjuvant, however nearly all participants showed traceable plasma levels of piperine. These traceable plasma levels of piperine were however nearly 10- to 60-fold lower than piperine plasma concentrations described in literature that show any substantial effect. Moreover, the results of our study could not confirm the results of the study by Shoba et al. where the bioavailability of curcumin increased by 2000% with addition of piperine (14). In this study 20 mg piperine was added to 2000 mg curcumin, which is comparable to the amounts of the supplements our participants have taken. Other herbal drugs or supplements were unlikely to have an effect on achieving therapeutically levels of curcumin.

In studies where β-glucuronidase was added prior to analysis, curcumin concentrations up to 3228.0 ± 1408.2 nmol/L were found. (24-26) The dosages of curcumin used in these studies were significantly higher than the advised dosage by the manufacturer. In our study, only one of the three participants using similar curcumin formulations was able to reach a curcumin C_max_ of 355 nmol/L. The difference between treatment with and without β-glucuronidase indicate that curcumin undergoes rapid and complete phase II metabolism. This, combined with the poor absorption, prevents therapeutic concentrations of curcumin. This is also observed in a pre-clinical study showing that after oral administration of 1.5 mg/kg curcumin formulated in a colloidal suspension (Theracurmin^®^) in Sprague-Dawley rats, unconjugated curcumin levels in the portal vein and abdominal portion of the vena cava were ranging respectively from 5.1±2.3 ng/mL and 2.1±0.9 ng/mL after 0.5h. Conjugated curcumin levels in the portal vein and abdominal portion of the vena cava were however 87.9±45.2 ng/mL and 65.1±46.3 ng/mL indicating a high first pass metabolism of curcumin. (27) One study in patients with pancreatic cancer and biliary tract cancer observed curcumin plasma levels of 324 ng/mL (range, 47-1029 ng/mL). Intravenous formulations could overcome the problems relating to the low bioavailability. These formulations will, however, be challenging due to the poor stability of curcumin in several solvents. (28) At neutral pH and a temperature of 37°C the t_1/2_ of curcumin is between 10 to 20 minutes. The t_1/2_ lowers by 50% in human blood incubated for 8 hours at 37°C. (28) The availability of curcumin will therefore become negligible due to these pharmacokinetics. Clinicaltrials.gov shows that liposomal curcumin is currently tested in clinical trials (NCT01403545, NCT04315350 (stopped inclusion due to COVID), NCT02369549), however, to date no results are reported. One dose-finding clinical trial (NCT02138955) did publish results, however of the 33 participants treatment was stopped in 23 patients after disease progression was found. In six patients the general medical condition deteriorated, with two early withdrawal and one study interruption. This study demonstrates that intravenous administration of liposomal curcumin is far from achieving any beneficial health results.

It is assumed that unconjugated curcumin is responsible for the therapeutic effect and that conjugation reduces the activity. (11) In several studies β-glucuronidase is added to the obtained plasma prior to analyses of blood samples. (20-22) Beta-glucuronidase hydrolysis glucuronidated and sulphated curcumin metabolites into unconjugated (free) curcumin. (29) These studies claim a higher plasma concentration of up to 185x unconjugated curcumin, while in fact this concentration is the sum of conjugated and unconjugated curcumin and the ratio between these two is not reported. (25). Nevertheless, these curcumin levels are assessed after the addition of β-glucuronidase. This leaves out the differentiation between conjugated and unconjugated curcumin. Consequently, no toxicity and no effect on NF-κB or pro-inflammatory cytokines was observed. (22)

Due to the nature of our study design we are able to establish that little to no systemic effect will be achieved with these curcumin plasma levels, however we cannot rule out any local effects in the lumen of the intestine that curcumin might induce. There are increasing reports of research in animals and humans where curcumin has been described having a potential beneficial effect on Inflammatory Bowel Diseases (IBD) like Crohn’s disease and Ulcerative Colitis. (30) It would therefore be more probable to expect beneficial health effects in local gut diseases than to expect a systemic health effect. Future research should assess this.

A limitation of our study is the use of β-glucuronidase to indicate whether conjugates of curcumin have been formed instead of directly quantifying these conjugates. However, this proves to be difficult due to the molecular properties of these conjugated compounds in our dual gradient mobile phase. A second limitation of our study is that by allowing the participants to bring their own curcumin formulation we introduced many variables like different dosages and a range of different formulations. Despite the fact that we let the participants take the curcumin in a controlled setting, we cannot guarantee that the curcumin formulations were of good quality. The goal of this explanatory study was however to assess curcumin plasma concentrations in everyday life use. Future research needs to be conducted to analyze curcumin in a randomized controlled clinical trial where different dosages and formulations can be analyzed.

## 5. Conclusion

In this observational trial, with patients and healthy participants using common available and home-made formulations of curcumin, non-therapeutic curcumin plasma levels were found. The quantified curcumin plasma concentrations were several orders below concentrations described to have a therapeutic effect in literature. The difference in conjugated and unconjugated curcumin can be observed by pretreating each sample with and without β-glucuronidase. Our results confirm that curcumin is rapidly and completely metabolized after oral intake. Several studies claim high curcumin concentrations, while they actually report total curcumin, including inactive metabolites. Currently available oral curcumin formulations are therefore still not sufficient in achieving systemic therapeutic concentrations. Future studies are needed to assess if patients with inflammatory bowel diseases may benefit from the effects of oral curcumin.

## Data Availability

All relevant data are within the manuscript and its Supporting Information files.

## Notes

### Competing Interest Statement

The authors have declared no competing interest.

### Clinical Trial

NL5931

### Funding Statement

The authors received no specific funding for this work.

### Author Declarations

The study protocol was reviewed and approved by the Ethics committee of the Amsterdam UMC, location Academic Medical Centre, The Netherlands.

